# Improving pandemic mitigation policies across communities through coupled dynamics of risk perception and infection

**DOI:** 10.1101/2020.07.09.20146985

**Authors:** Matthew J Silk, Simon Carrignon, R. Alexander Bentley, Nina H Fefferman

## Abstract

Decisions to adhere to health-protective behaviors (e.g. mask-wearing, social distancing, etc.) that impact the spread of COVID-19 are not made in isolation by each individual. They are instead the result of the social construction of perceived risks and resulting community norms. In populations in which disease is unlikely to spread throughout all communities simultaneously, community-driven perception of risk can drastically alter collective outcomes. A community could respond to a few infections by becoming fearful and adopting anticipatory behaviors that protect them from disease spread. Similarly, there could be false reassurance, in which low disease incidence over time leads to community consensus that protective actions are unnecessary (even if they are the reason cases remains rare). We therefore model COVID-19 spread with three synergistic dynamics governing individual behavioral choices: (1) Social construction of concern, (2) Awareness of disease incidence, and (3) Reassurance by lack of disease. We use a multiplex network approach that captures social communication and epidemiological spread. We find that effective protective policies enacted too early may backfire by allowing a community to become reassured and therefore unwilling to adopt or maintain further protective behaviors. Based on these insights, we propose that public health policies for which success relies on collective action should be designed to exploit the *behaviourally receptive phase*; the period between the generation of sufficient concern as to foster adoption of novel protective behaviors and the relaxation of adherence driven by reassurance fostered by avoidance of negative outcomes over time.

## INTRODUCTION

The ongoing COVID-19 pandemic has cemented the understanding among public health researchers, practitioners, and policy makers that the spread of infectious disease is more than a purely epidemiological process. While COVID-19 has strained hospital capacity (1), the global supply of personal protective equipment (2, 3), food supply chains (4, 5) and unemployment insurance processing capacity (6), the greatest challenges in understanding, predicting, and planning mitigation for the ongoing spread of the disease lies in how to understand, anticipate, and influence human behavioral responses (7).

Efforts to incorporate social (8), psychological (9), or economic (10) factors have revealed the profound effects of behavioral choices on projected outbreak dynamics (11, 12). Critically, however, many studies helping shape policy have considered behavioral factors as mostly uniform across the affected population and mostly constant throughout the course of an outbreak (13, 14). While there are legitimate and important reasons to explore models making these assumptions, they do not reflect the current reality of behavioral responses to the COVID-19 pandemic.

People’s behavioural responses to the pandemic will vary considerably, often between locations and over time, driven by variations in local government policy as well as individual behaviors over time (e.g. as “stay at home” orders were enacted and relaxed; (15)), and across social and demographic groups (e.g. conservative vs. liberal; (12, 16)). These are not static parameters; these shifting patterns of behavior are not independent of the spread of infection but are instead inextricably coupled with the spatial and temporal patterns of disease incidence.

Models that have incorporated individual behavioral responses with the epidemiological dynamics of an ongoing outbreak (e.g. avoiding sick people, accepting a vaccine; (17-23)) provide the basis for exploring more dynamic models that allow individual behaviors to change when some psychological threshold for change is met. Understanding, analyzing, and predicting such coupled dynamics, in which behavioral responses themselves shift over space and time, will allow us to build outbreak models that can apply broadly across regions and withstand shifting conditions for more than a few weeks at a time. Such models can then support policy makers in designing flexible, responsive plans that can be better communicated to, and accepted by, the public, improving efforts to mitigate current risks and helping us prepare for future pandemics.

Amenable to this dynamic behavioral approach, network models in epidemiology have been used with great success to understand how individual heterogeneity in contacts among individuals can cause deviations in outbreak progression relative to homogeneous, mean-field approximations of average behaviors (24-27). Analogously, network models of “social contagions” predict the spread of beliefs or information through populations (28-33). Studies focusing on specific aspects of social contact networks have shown that a variety of structural features — edge density, clustering coefficient, modularity — of those networks affect the progression of epidemics and information (34-40).

For social-behavioural phenomena, multiplex (also called multilayer) networks capture ongoing, coupled dynamics [e.g. (41-43)]. In application to coupled behavior-epidemiological dynamics, here we use a multiplex network to combine a physical contact, or “infection,” network over which an infectious disease might be transmitted and a communication network over which information or opinions might be shared can therefore provide the required tool for coupling the states and dynamics between the layers (44, 45). Consequently, simulations exploiting coupled multiplex networks have provided important insights into the impact of social construction of risk perception on the spread of infectious diseases [e.g. refs (46, 47)].

Social distancing exemplifies how coupling between a communication and infection network layer is critical to the COVID-19 pandemic. Adherence to social distancing recommendations is determined solely by individual concern and resulting behaviors. Those concerns are, in turn, constructed by each individual based both on direct observation of evidence (e.g. contact or lack of contact with sick people), and on social inputs, including communication with worried peers/advisors and perception of social norms (e.g. knowledge of the compliance of others). These social inputs rely on a communication network which may or may not overlap with the contact network over which infection spreads. The communication network and contact network can be considered as layers of a multiplex network containing the same set of interacting individuals. Coupled dynamics between the communication and infection layers of the multiplex network can have some profound impacts on the progression of epidemics and the efficacy of attempted mitigation strategies. Characteristics of the communication layer will impact which individuals and communities perceive risk to be high (whether or not the actual risk of infection is high in their physical environment), which will influence rates of adherence to disease-defensive public health policies. For the same reasons, multiplex structure should influence when disease permeates heterogeneous networks, reaching different communities at different time/stages of the outbreak, and how rapidly it spreads through each community when it does reach them, especially as increased disease prevalence should increase perceived risk, and thereby slow the progression via behavioral defensive responses.

Two of the most likely network features to be important in shaping the interplay between these dynamics are homophily (the tendency to affiliated with similar others) (48-51) and modularity (the strength of division between “communities” ; populations more likely to be connected to each other than to individuals outside of the group) (39, 52, 53). To begin to understand how these coupled behavioral-epidemiological dynamics may be driving the current COVID-19 pandemic, we here present a multiplex network model that captures a standard Susceptible-Exposed-Infected-Recovered (SEIR) epidemiological dynamic (54), in a population with a simplified demography (children, adult, and elderly), and with two opposing “predispositions” that contribute to homophily in either the infection layer of the network, communication layer, or both. These predispositions may also determine how easily someone becomes alarmed about personal disease risks (factors in predisposition could include political or religious beliefs, personality traits, or socioeconomic status). Networks are organized into social communities, with the modularity of these subdivisions also able to differ between the contact and communication layers of the network. We assess the severity of disease progression by a joint measure considering both the time and magnitude of the epidemic peak in each community (see details in Methods) to allow for discussion of public health strategies that focus both on sufficient time to act and have the capacity for action to handle peak surge demands.

Our models assume that social influence can be effective at elevating individual concern about disease, but only direct observation of an absence of infection in a community will provide reassurance (i.e. cause a decrease of concern). We use this model to explore the impact of modularity and homophily according to predisposition regarding the severity of disease progression through communities, the relative impact of social vs. observational contributions towards estimation of disease risk on the disease outcomes in communities, and the impact of reassurance on the likely dynamics in risk behaviors over time, again affecting disease outcomes. While this model is parameterized to reflect current COVID-19 features and challenges, it seems clear that socio-behavioral dynamics that shape the nature of risk perception, and therefore disease-defensive behaviors should be important for any pandemic preparedness planning for the future. We therefore here explore and present a broad set of scenarios.

## RESULTS

We simulated the spread of infection and concern through nine multiplex networks that differed in community structure and homophily between two predispositions (see Fig. 1 for an example). An individual’s concern about infection determined their probability of social distancing. Individuals that socially-distanced cut 50% of their social contacts in the infection layer, reducing their chances of becoming infected and helping to “flatten the curve” (Fig. S1). Concern could change over time through three learning processes: a) Social Construction of Concern, in which an individual’s concern increased depending on the proportion of their connections in the communication layer of their network who were adherent to social distancing; b) Awareness, in which individuals became more concerned based on each person they knew in the communication layer of their network who was sick; c) Reassurance, in which individuals concern decreased at every time-step at which none of their connections in the communication layer of the network were ill. We monitored the epidemiological outcomes and changes in adherence to social distancing over time separately for each of 10 communities or modules in our social networks (see Fig. 1).

**Figure 1.**
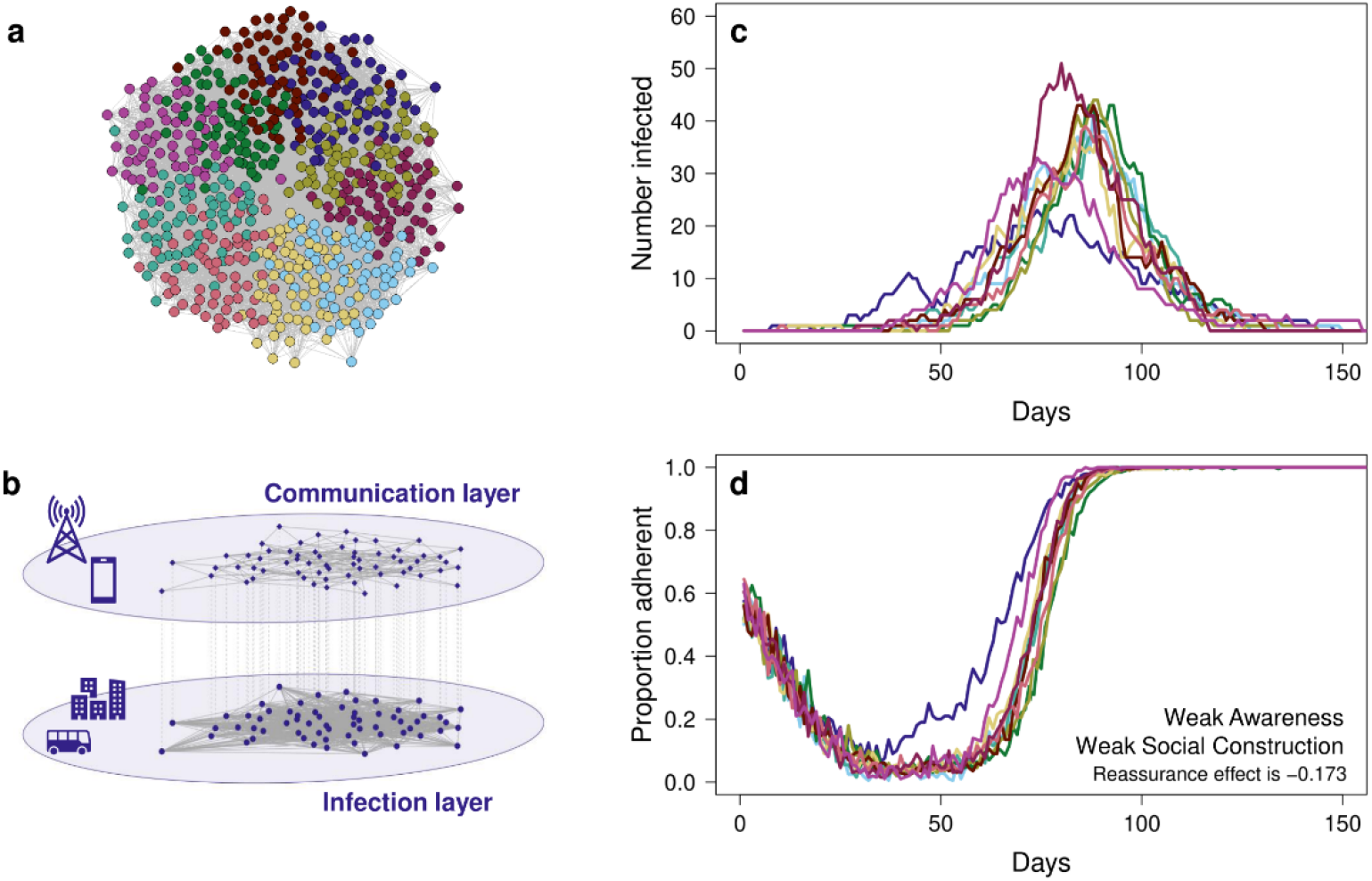
A depiction of the modelling process. We generated coupled multiplex networks (a, b) containing a communication and infection layer for 2000 individuals. This network was divided into 10 equally sized social communities and the network was rewired to have a desired modularity (a). Here (a) we show community assignment in the infection layer for young adults of a single predisposition (i.e. only part of the network). b) The infection layer and communication layer of the multiplex network differed in density so that the infection layer was better connected and could have the same or higher modularity and the same or lower homophily according to predisposition. We then modelled the spread of infection (c) and change in adherence to social-distancing (d) and recorded these at a community-level. The colours of the lines depict the community membership illustrated in (a).

### Communities infected later are often hit harder

In the absence of any form of learning, communities hit later in the epidemic typically have more severe outbreaks than those hit earlier (Figs. 2 & S2, Tables S1-5), with a reduced time from the start of an outbreak to its peak, and a higher peak (Fig. S1). This pattern remains regardless of whether initial social distancing levels are intermediate (50% of the population adherent; Table S1) or low (20% adherent; Table S2). These patterns were qualitatively similar across all of the nine multiplex networks simulated. Outbreaks were typically more severe, but with severity increasing less over time, when the infection layer was less modular. Outbreaks were also more severe on average when modularity in the infection layer and communication layer were mismatched (i.e. the infection layer was more modular than the communication layer), except in the case when there was matching homophily in connections in both layers (Tables S1-S5).

**Figure 2.**
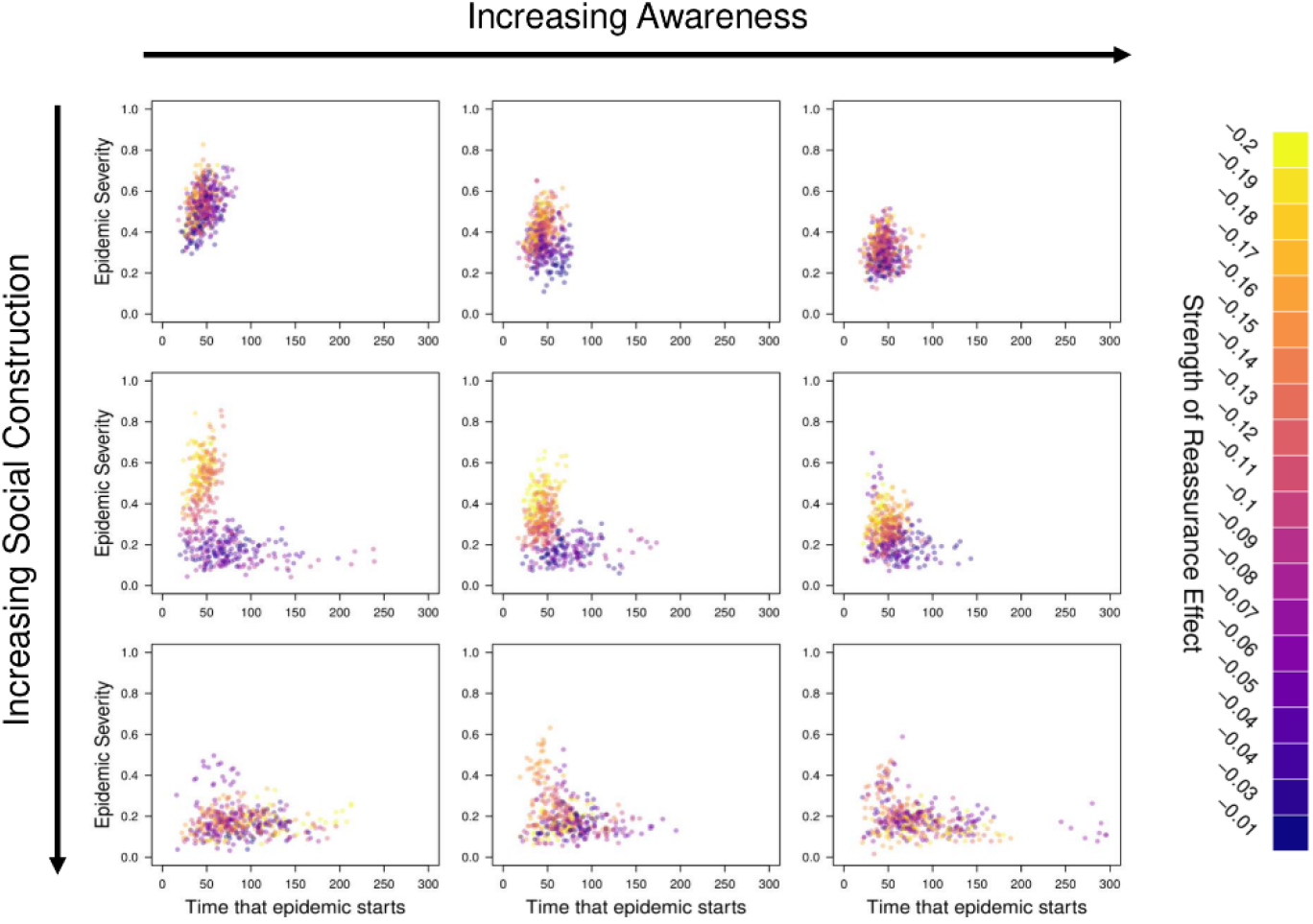
The relationship between the time that an epidemic starts in a community and the severity of that epidemic for a range of different Awareness effects (0, 0.4, 1.6) and Social Construction effects (0, 0.2, 1). In the top row there is no change in the concern of individuals through social construction, while in the left-hand column there is no effect of awareness. Conversely these two parameters take their maximum values in the bottom row and right-hand column respectively. Points are coloured on a continuous scale according to the strength of the Reassurance effect. For each run of the simulation this was sampled from a uniform distribution between -0.2 and -0.01.

### Different forms of learning flatten the curve in different ways

Increased adherence to social distancing through Awareness of ill individuals and through Social Construction of concern both reduced the severity of the outbreak but also resulted in strikingly different epidemic outcomes (Fig. 2). When individuals increased social distancing due to Awareness, the height of the epidemic peak was reduced but the time to reach the peak was not (Fig. 2). In contrast when individuals became more likely to be adherent to social distancing through Social Construction both the height of the epidemic peak and the length of time between the start of an outbreak and epidemic peak increased. This resulted in the timing of epidemic peaks becoming more variable between communities (Fig. S3) which may have important implications for the mitigation.

Social Construction of concern had a much greater impact on mitigating epidemic severity than awareness of actual incidence of illness among those with whom they communicate. Only moderate influence of the social norms of communication neighbours had a sizeable mitigating effect. It is important to note, however, that low levels of Social Construction were ineffective (Fig. 2). In contrast, increases in the impact of Awareness had a more linear effect on the outcome of outbreaks (Fig. 2).

Results were qualitatively similar regardless of the modularity or homophily of the network. However, the importance of Social Construction of concern in mitigating outbreak severity was highest when the modularity of the communication network was lower (Q_rel_=0.4) and when there was homophily according to predisposition (Tables S1-5).

### Social Construction of concern driving the probability of adherence to social distancing is particularly important for later hit communities

Increased adherence through Social Construction, but not as a result of awareness, helped later-hit communities disproportionately whereby, rather than being more severe, outbreaks in these communities were no worse or even less severe than those in communities hit early (Tables S1-2). Similarly, to the previous result, this outcome was not achieved with only weak Social Construction (Tables S1-2). The importance of Social Construction in mitigating this effect was highest when the infection layer of the multiplex network was more modular (Q_rel_=0.6) and in its absence there was the steepest increase in outbreak severity with later starting date (Tables S1-5).

### The Reassurance effect when neighbors in the communication layer are all healthy amplifies these differences

In the absence of Social Construction, the reassurance effect of people becoming more relaxed in their probability to adhere to social distancing over time is critical for epidemic dynamics. When Social Construction of concern is absent, or only increases concern slowly, then a strong Reassurance effect causes outbreaks within communities to be more severe on average and disproportionately impacts later-hit communities so that outbreaks tend to be much more severe (Fig. 3a). It also reduces variability in outbreak peaks between communities (Fig. S4) resulting in greater population-level synchrony in epidemic dynamics.

**Figure 3.**
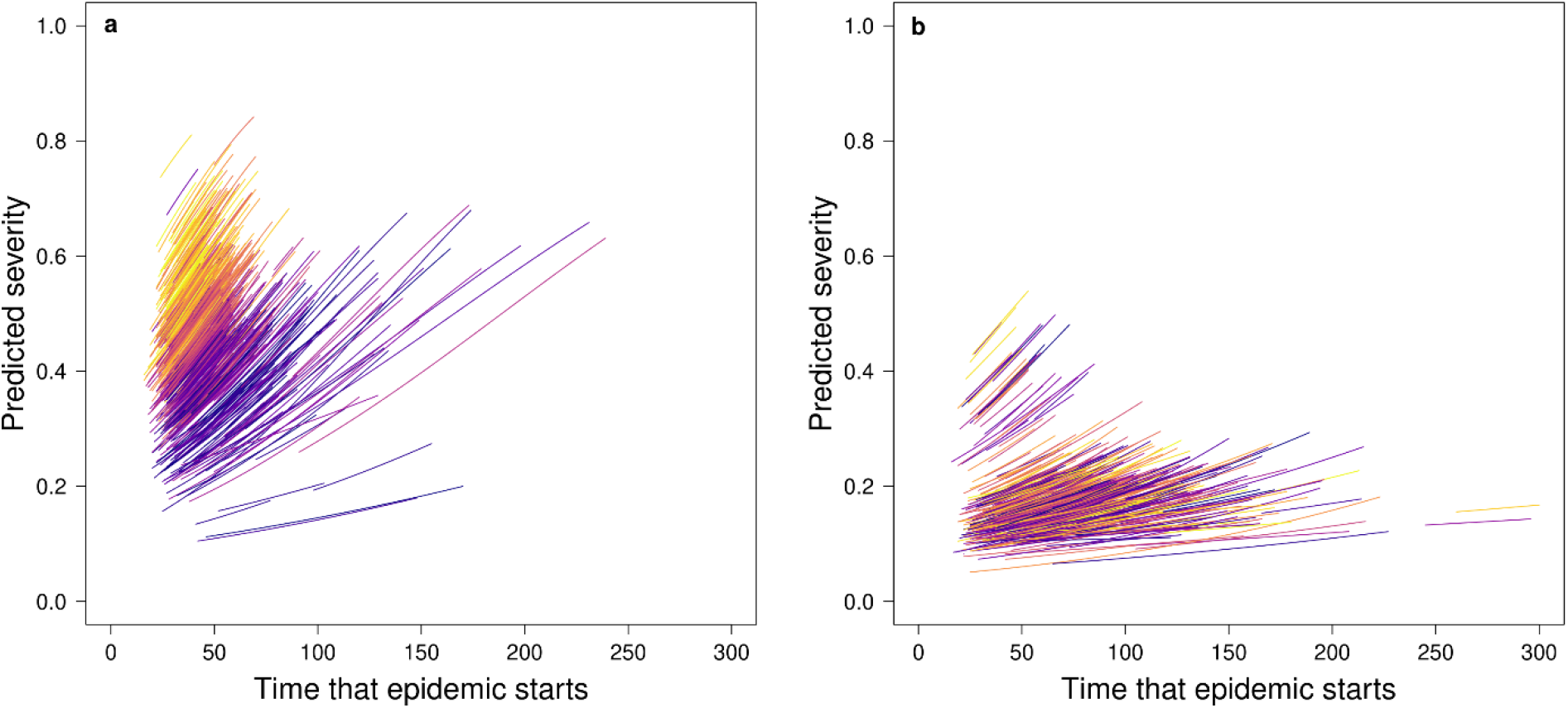
Random effect estimates for the effect of each simulation run from the statistical model fitted to explain epidemic severity in the absence of a Reassurance effect. We show lines calculated from point estimates for the random intercept and random slope for when a) Social Construction is weak (0, 0.1, 0.2) and b) Social Construction is strong (0.5, 1). Lines are coloured on a continuous scale according to the strength of the Reassurance effect. For each run of the simulation this was sampled from a uniform distribution between -0.2 and -0.01. Yellow indicates a strong reassurance effect and purple indicates a weak reassurance effect. For the purposes of plotting the lines we assume the Social Construction effect is fixed at 0.1 (low) for a) and fixed at 1 (high) for b). We assume no Awareness effect and use estimates that apply to Network ID 1 (modularity of 0.4 for both layers and no homophily according to predisposition).

However, when Social Construction is stronger, the strength of the Reassurance effect is much less important in explaining outbreak severity (Fig. 3b), with similar epidemic outcomes even if it is assumed that the tendency for people to be adherent would otherwise decline quickly over time in the absence of knowing anyone who is sick.

This result is caused by there being a very strong relationship between outbreak severity and the minimum proportion of each community adherent to social distancing (Figs. 4 & S5). When people pay more attention to the concern of their network neighbours more, the proportion of non-adherent people remains low even in later-hit communities (Figs. 4 & S5). As above, the effectiveness of Social Construction of concern in preventing the adverse effects of relaxation over time has a steep threshold, meaning that maintaining the importance of social norms throughout an outbreak is important.

**Figure 4.**
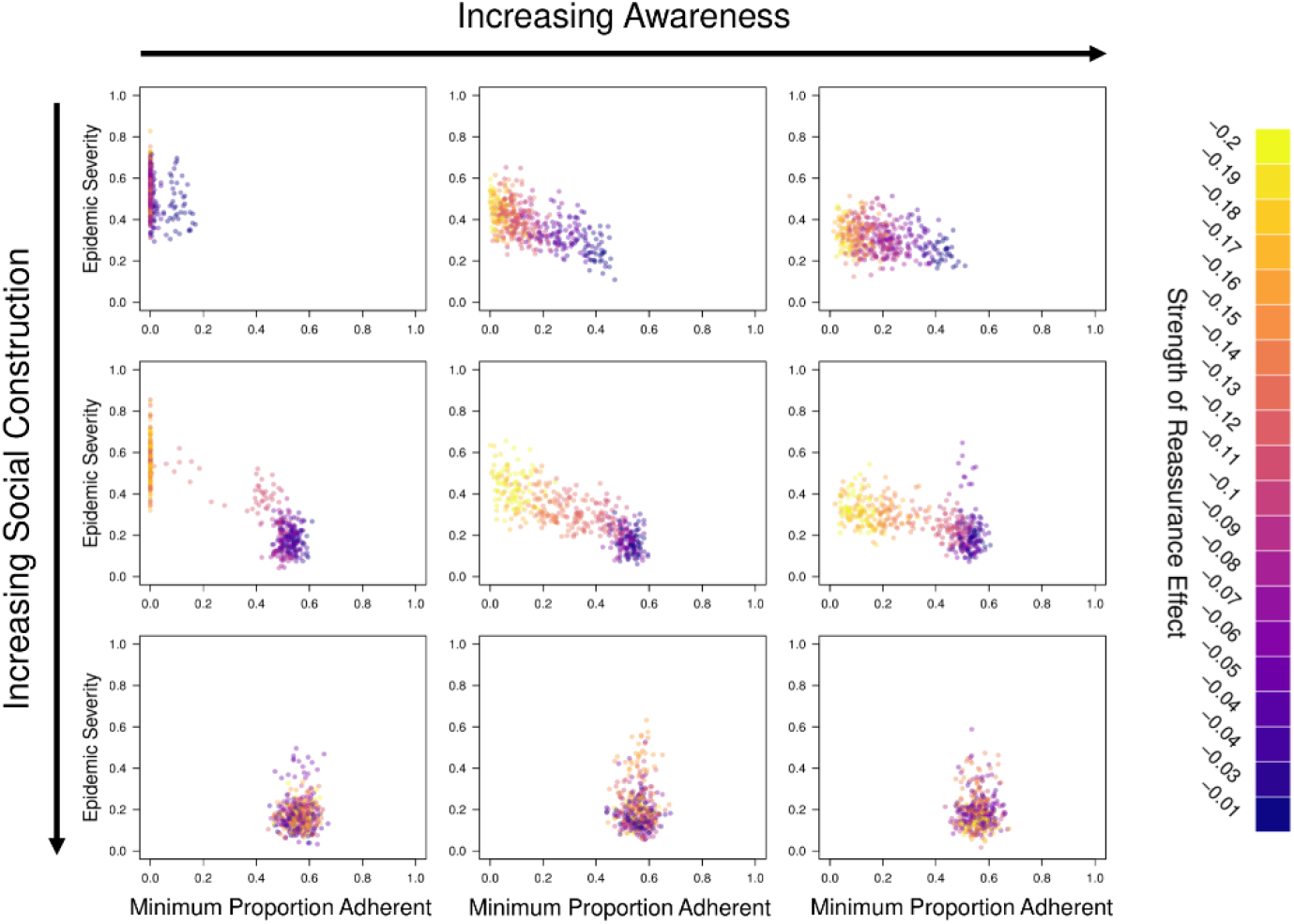
The relationship between the minimum proportion of people adherent with social distancing in a community and the severity of that epidemic for a range of different Awareness effects (0, 0.4, 1.6) and Social Construction effects (0, 0.2, 1). In the top row there is no change in the concern of individuals through Social Construction, while in the left-hand column there is no effect of Awareness. Conversely these two parameters take their maximum values in the bottom row and right-hand column respectively. Points are coloured on a continuous scale according to the strength of the Reassurance effect. For each run of the simulation this was sampled from a uniform distribution between -0.2 and -0.01

After controlling for other factors, the negative correlation between the strength of the Reassurance effect and both outbreak severity and the increase in outbreak severity over time was relatively consistent between the nine networks modelled (Figs. S6-7). The negative correlation was strongest when the infection layer was more modular than the communication layer and there was homophily only in the communication layer resulting in an important mismatch between the structure of the two layers.

## DISCUSSION

Our results clearly show the importance of all of the considered mechanisms (Awareness of infection, Social Construction and Reassurance) in driving concern and shaping the ongoing dynamics of disease progression (in both timing and severity) through distinct social communities. Understanding how each mechanism acts, both independently and synergistically with the other mechanisms, will be of critical importance in accurately anticipating infection risks in large, heterogeneous populations. Differences in concern about risk between communities will clearly amplify patchiness in spatiotemporal spread of infection (Fig. 2). One clear, unfortunate impact of effective protective behaviors that serve to buffer or insulate a community from early disease spread is that they can foster social reassurance, degrading concern in the need to maintain them (Fig. 3a). It may therefore be that communities hit later in the progression of disease spread throughout the entire population may be paradoxically less well aligned towards behavioral defenses than if those communities had experienced greater numbers of infections earlier on in the outbreak.

For the spread of COVID-19, these results indicate the potential for a “perfect storm”. The delay between behavioral response and local increase in disease prevalence hinders concern-based protective behaviors, whether based on observational awareness or on Social Construction. The infection network structure of the United States is itself a modular patchwork centered around towns and cities of vastly different population sizes (55). The community structure of communication for individuals within and between these municipalities is also clearly highly modular and has been shown highly homophilic in its belief structures on topics correlated to behavioral decisions regarding infection risks during COVID-19 (49). This creates patchy echo boxes of communities that are temporally and spatially insulated from experiencing the same risks at the same times, even while centralized media reporting and early scientific investigations discussed COVID-19 risks as nation-wide problems.

Policy may have therefore inadvertently exacerbated frustration by enforcing protective behaviors in populations that had not internalized social norms of increased concern, nor had yet experienced sufficiently high levels of infection as to be observable by the average individual. In this way, protective policies likely served to insulate these communities from the initiation of widespread transmission of disease, slowing the timeline for observable infection, and thereby allowing the Reassurance effect to *decrease* in concern, and therefore in the belief in the need for action. In a very practical sense, the jarringly imposed adoption of “shelter at home” protocols in regions without widespread outbreaks likely allowed for the normalisation of perceived risk of infection before the actual escalation of local disease incidence could be felt, leading to rejection of the normalisation of the protective behaviors. If correct, we are just now primed by these seemingly benevolent actions to experience worse outbreaks in thus-far-minimally-affected areas that should have been able to use the delay to plan and prepare (both practically and psychologically) to more effectively mount mitigation efforts when disease inevitably arrives.

Importantly, these models allow us to move beyond trying to explain currently observed patterns or predict near-term coupled behavioral-epidemiological dynamics across communities. They also allow us to consider shaping policies that rely on the normalization of risk mitigation behaviors when the population may be most accepting of, and therefore adherent to them due to having already normalized perceived risk of infection. By timing and scaling the magnitude of proposed interventions to match both individually observable disease incidence and the socially-constructed concern for disease risks in each community, we may actually be able to prevent spread more effectively than enforcing draconian measures that would be more epidemiologically effective with broad public adherence, but are also more and more likely to be rejected over time. Dynamic protective policies are not novel. Models based only on epidemiology and healthcare capacity have already proposed pulsed strategies for “shelter in place” orders, in some cases suggesting that these pulses could continue even until a vaccine enables us to achieve herd-immunity levels of protection (14). Instead, we propose that such dynamic models should pulse in accordance with community-level concern. Critically, we do not mean to suggest that mitigation efforts should respond/react to concern-driven demand, but instead that policies can anticipate when they might be well enough received to enable concern-driven acceptance and adherence. Ultimately, even an ideal public health protocol will be compromised if it does not achieve support and adherence from the public it strives to protect. Vaccination is an excellent example – MMR inoculation can eradicate regional measles, mumps, and rubella risks, but outbreaks in highly developed and resource-rich nations still occur due to parental refusal of vaccination. Public health efforts now focus on increasing adoption rather than improving the vaccine or deliver of the vaccine. We propose that policy for behavioral defensive actions (which are currently the only effective means we have for mitigating COVID-19, though vaccines and therapeutics are actively being researched) should follow in the path of public health policy for vaccination and incorporate explicit focus (including modeling efforts) to anticipate public concern, acceptance of, and adherence to, any recommendations.

Beyond the results presented here, there are obvious, immediate, additional questions that need to be considered. These range from characterizing regional differences in the overlap between communication and infection network layers for average individuals, through experimenting to understand how different relative impacts of contributions to each of the three mechanisms of learning contribute to individual behavior over time, to extending beyond these three initially considered learning mechanisms themselves. Perhaps most importantly, these models have ignored the role of fully asymptomatic infections (i.e. those who never develop symptoms of disease) in driving spread in the disease layer without a concomitant impact in the communication layer. If (as is currently suspected but not yet known (56)), the probability of remaining fully asymptomatic is partially age-dependent, then communication neighborhoods among similarly aged individuals may exacerbate differences between community-driven risk perception with different age distributions simply by this failure of infections to be detected. These effects may go far beyond the simple community dynamics we have investigated here. That said, even from just these first models, there are already clear policy implications.

Strong, reinforced social norms are critical in maintaining a community’s adherence to social distancing over time. It is likely that there is a highly sensitive window during which the perception of risk is high enough to foster adoption of protective behaviors that may be normalized before lack of global prevalence provides false reassurance that could undermine their adoption. Once normalized, behaviors may be passively (rather than actively maintained), meaning that reassurance is less likely to decrease their observance. We call this window the *behaviourally receptive phase*. Reassurance effects easily compromise effective early interventions, even in the absence of social amplification (social construction could only increase concern in our models), meaning that communities that succeed in delaying incipient outbreaks may experience worse overall outcomes than those hit with higher incidence earlier on. Beyond understanding how and why these dynamics might be observed, these models also suggest methods for purposeful intervention based explicitly on combatting reassurance effects. Asking community leaders to be vocal about their understanding of risk in the community can support ongoing social construction of concern that will strengthen social norms around protective behaviors. Municipal efforts to support families with loved ones who have been diagnosed may meaningfully amplify awareness in ways that combat reassurance while focusing on positive aspects of community rather than individual fear and isolation.

Our work has demonstrated the critical and inextricably intertwined roles that social construction of risk perception, community awareness of disease incidence, and reassurance effects from local absence of active cases can have on the success of outbreak mitigative policies. We see footprints of these effects in the observed dynamics of the COVID-19 pandemic as we write, and we advocate for additional discussion building on these explicit insights to become a greater focus for both research and policy.

## METHODS

### Overview

We used stochastic simulations to test how the spread of concern influenced epidemic dynamic of Covid-19 through populations divided into communities in both communication and physical contact structures. All simulated populations consisted of 2000 individuals of three age-categories: children, young (low-risk) adults and old (high-risk) adults and split between two distinct “predispositions”. We then simulated realistic social networks for our populations, using coupled (multiplex) networks for the spread of infection and the spread of concern about the disease (Fig. 5). We recorded epidemic outcomes for a range of network structures and proposed models for the change in concern of individuals over time. All modelling was conducted in R3.6.1 (57). All R code used is provided in the Supplementary Information and on GitHub (https://github.com/matthewsilk/CoupledDynamicsNetworkPaper/). Full methods are detailed in the Supplementary Material.

**Figure 5.**
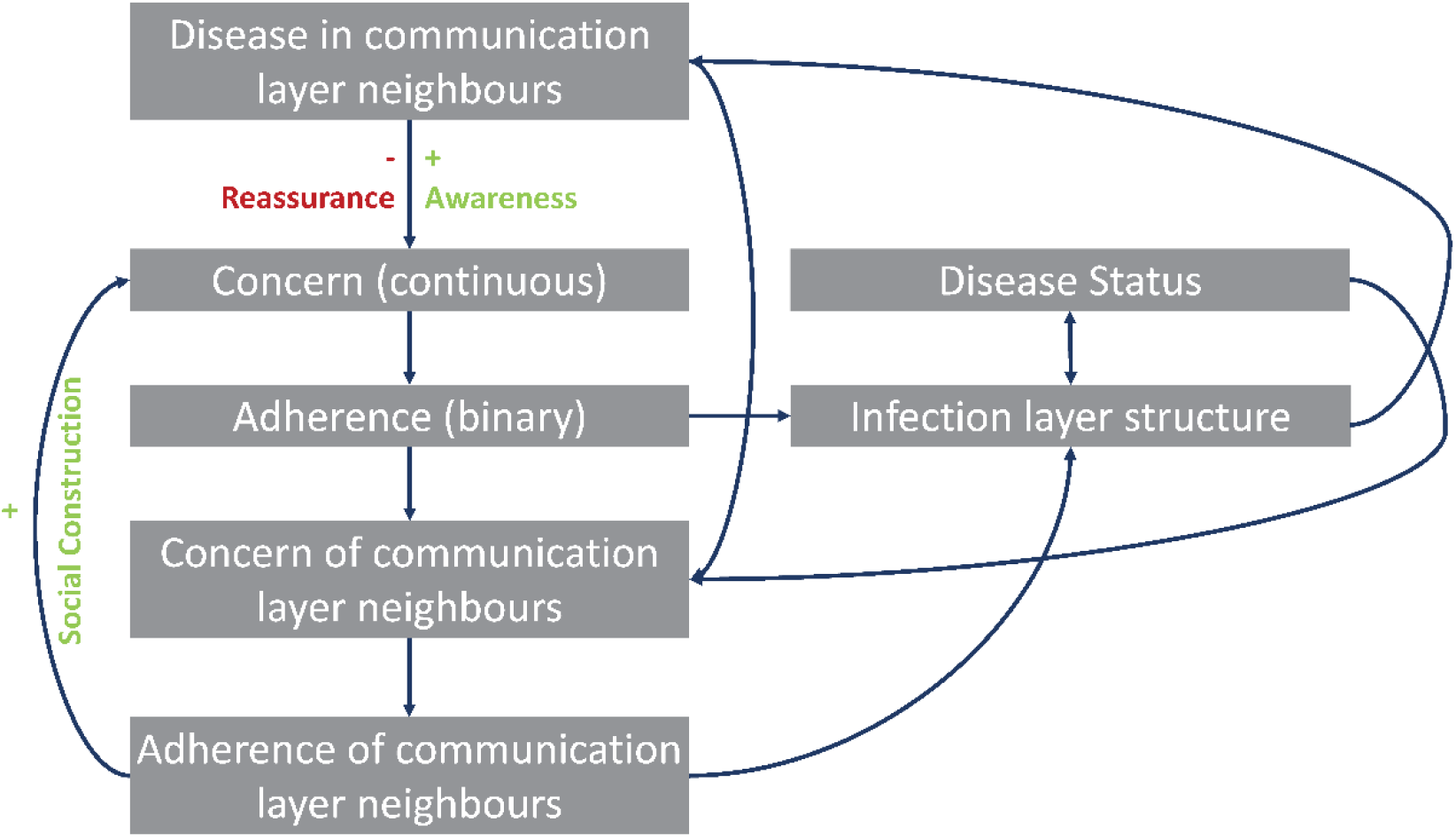
A schematic providing an overview of how our stochastic model couples risk perception and infection dynamics

### Population generation

Our population was 24% children, 63% young adult and 13% old adult to match recent US demographic data. Age classes could differ in the social connections, epidemiological outcomes and concern about the disease (as detailed in relevant sections). Individuals also had one of two baseline predispositions: “A” and “B”. 50% of individuals were of each predisposition. Homophily according to predisposition could impact social connections.

### Social network generation

We generated 9 multiplex social networks that connected all individuals within a “communication” layer that influenced the spread of concern about the disease and an “infection” layer that influenced the transmission of the pathogen itself. Networks were simulated in a stepwise fashion so that connections in both the communication layer and infection layer displayed community structure, homophily by age and could display homophily by predisposition (see Supplementary Methods). Children were connected to parents in the young adult layer and shared their parents’ connections with older adults. We used three different combinations of modularity and three different combinations of homophily by predisposition. Using relative modularity (53), our networks either had a modularity of 0.4 in both layers a modularity of 0.6 in both layers, or the infection layer had a modularity of 0.6 and the communication layer a modularity of 0.4 (to reflect the fact that more communication is likely between communities than contacts relevant for infection). The proportion of each predisposition (A and B) within each community was the same as that in the overall population. For homophily by predisposition we included networks in which there was either a) no homophily in either layer, b) homophily in the communication layer only, or c) homophily in both layers.

### Concern model

We modelled the spread of concern about the disease through the communication layer as a complex contagion (58). Whether an individual was adherent or not (binary trait) depended on a Bernoulli draw based on an underlying probability which we term concern. Consequently, it was possible for individuals to move from being non-adherent to adherent but also for them to return to being non-adherent. Individuals with intermediate levels of concern were likely to fluctuate between adherent and non-adherent states. Initial levels of concern resulted in a 50%, 20% or5% chance of adherence. The underlying concern of all adults could then be influenced by Social Construction, Awareness or Reassurance (Fig. 5).

Each time they became adherent, individuals cut their connections within the infection layer of the network while maintaining their connectivity in the communication layer. Individuals cut connections with a 50% probability (designed to reflect a reasonable approximation of a real-life effect), with connections being cut to have an edge weights of 0.001 meaning that the probability of transmission across them was negligible. If an individual became non-adherent then these edge weights returned to their initial values.

### Infectious disease model

We modelled the spread of SARS-CoV-2 using an age-structured stochastic SEIRD model adapted from (54) as detailed in the Supplementary Methods. The model contains susceptible (S), exposed (E), pre-symptomatic (I1), symptomatic (I2), hospitalised (I3), recovered (R) and dead (D) compartments. The transition between compartments is detailed below and parameter values are provided in Table S6. Symptomatic (I2) and hospitlalised (I3) individuals cut all connections in the infection layer of the network to 0.001. These connections were restored to their full value if and when individuals recovered.

### Simulations

For each of the 9 multiplex networks studies we conducted simulations for the full combination of starting values (3), social construction effects (5) and awareness effects (5). For each of these 75 combinations we conducted 5 replicate simulations with the same network structure but with different individuals seeded with infection. A unique value of the reassurance effect was drawn from the uniform distribution defined above for each run of the simulation. We simulated a time period of 300 days (or until there were no remaining infected individuals). The simulation algorithm is detailed in the Supplementary Methods.

### Analysis

To test the combined effects of social construction of concern, increases in concern due to awareness of incidence of disease, and relaxation of adherence to social distancing due to reassurance effects from not knowing infected people, we first calculated a measure of outbreak severity that took into account the height and timing of the epidemic peak in each community (see Supplementary Methods). We then used linear mixed effects models to ascribe variation in outbreak severity to network structure and values of the Social Construction, Awareness and Reassurance effects as detailed in the Supplementary Methods.

## Data Availability

NA

https://github.com/matthewsilk/CoupledDynamicsNetworkPaper/

## Acknowledgments

We thank Mario Small for useful conversations in formulating these questions and for comments on the draft. We thank Dave Hodgson for allowing MJS time to pursue this work. We also gratefully acknowledge funding support for this work from NSF DEB 2028710.

